# Sex Differences in the Impact of Childhood Socioeconomic Status on Immune Function

**DOI:** 10.1101/2020.09.30.20204925

**Authors:** Jeffrey Gassen, Jordon D. White, Julia L. Peterman, Summer Mengelkoch, Randi P. Proffitt Leyva, Marjorie L. Prokosch, Micah J. Eimerbrink, Kelly Brice, Dennis J. Cheek, Gary W. Boehm, Sarah E. Hill

## Abstract

We examined relationships between multiple sources of early life stress and adult immune function in humans. Adult participants provided retrospective information about their childhood a) socioeconomic status, b) household unpredictability, and c) exposure to adverse experiences. Participants’ peripheral blood mononuclear cells were then isolated for use in functional assays: a) tumor cell lysis by natural killer cells, and b) phagocytosis of *Escherichia coli* bioparticles, and c) mitogen-induced leukocyte proliferation and cytokine release. In men, lower childhood socioeconomic status predicted decrements in immunological performance across functional assays, along with greater spontaneous cytokine release from PBMCs. These changes co-occurred with elevations in plasma testosterone levels. Similar effects were not observed for other sources of stress, nor were they found in women (with the exception of spontaneous cytokine release). These findings provide evidence that low childhood socioeconomic status has a lasting negative impact on multiple aspects of immune function, particularly in men.

---

Over the last three decades, a considerable body of research has demonstrated that individuals exposed to stressors during childhood exhibit an increased risk for a number of chronic health problems later in life, including metabolic disorders (Delpierre et al., 2018), cardiovascular disease (Murphy, Cohn, & Loria, 2017), mental health problems (Mock & Arai, 2011), and susceptibility to infectious illnesses (Cohen et al., 2004). This propensity is found to operate independently of an individual’s adult economic circumstances (Galobardes, Lynch, & Davey Smith, 2004; Kittleson e al., 2006; Taylor, 2010), suggesting that the impact of early life environments on health is long-lasting and not easily reversed. For example, one study found that exposure to childhood socioeconomic disadvantage predicted an increased risk of cardiovascular disease in a cohort of male physicians, despite their achieved adult socioeconomic status far exceeding that of the average American (Kittleson et al., 2006).

Despite the well-established link between childhood conditions and adult health (Delpierre et al., 2018; Mock & Arai, 2011; Murphy et al., 2017; Taylor, 2010), the myriad environmental and biological pathways through which one’s early life experiences impact adult health outcomes are only beginning to be understood. Initial clues about contributors to this relationship have surfaced in research demonstrating a link between early life adversity and the tendency to exhibit an exaggerated inflammatory response to immunological stimulation (Levine, Cole & Weir, 2015; Miller & Chen, 2010; Miller et al., 2019). Additional clues are found in research demonstrating that adults who report lower childhood socioeconomic status (SES) exhibit increased susceptibility to an experimentally-induced viral infection relative to those reporting higher childhood SES (Cohen et al., 2004). However, little is known about the broader immunological context in which such tendencies emerge (i.e., which specific facets of immunity are impacted by early life disadvantage) or the extent to which individual-level factors buffer or exacerbate the impact of adverse childhood environments.

One individual-level factor that is likely to moderate the relationship between early life environments and immunological function is biological sex. Indeed, although much research finds that developmental experiences can have a lasting impact on both men’s and women’s health (Delpierre et al., 2018; Mock & Arai, 2011; Murphy et al., 2017; Taylor, 2010), many studies find these effects to be greater for men than women (Aiken & Ozanne, 2013; Del Giudice et al., 2018; Geary, 2016; Stinson, 1985; for exceptions see Gallo et al., 2018; Laceulle et al., 2014). For example, early life exposure to infection and nutritional stress each have an asymmetrically negative impact on men’s physical development and health compared to women’s (e.g., Kuzawa & Adair, 2003; Stinson, 1985; Yap et al., 2012). Additionally, intervention programs aimed at improving health in the context of poverty are often disproportionately beneficial for boys compared to girls (e.g., Campbell et al., 2014; Conti, Heckman, & Pinto, 2015; Garcia et al., 2016). Such results suggest greater health-related developmental plasticity in males compared to females, with male health being more susceptible to environmental influences than female health, for better and for worse.

Existing explanations for why men are often more susceptible to the health consequences of early life stress than women include factors such as sex differences in hormone levels (e.g., androgens; Del Giudice et al., 2018), stronger canalization of bodily growth in women than men (Stinson, 1985), and a greater tendency for men to engage in health-harming behaviors in stressful environments compared to women (Mustard & Etches, 2003). Research in the evolutionary sciences suggests that such differences reflect sex differentiated energetic investments in mating effort vs. somatic development and repair (Brumbach et al., 2009; Del Giudice, Gangestad, & Kaplan, 2015; Nunn et al., 2009; Rolff, 2003; Stearns, 1992; Stoehr & Kokko, 2006). According to this perspective, males in stressful ecologies should upregulate investment in mating effort (at the expense of downregulating investment in somatic development / repair) to minimize the likelihood of perishing without having successfully reproduced (Belsky, Schlomer, & Ellis, 2012; Brumbach et al., 2009; Ellis et al., 2009). This pattern is predicted to occur in males but not females because females’ ability to reproduce is highly dependent on physical condition (especially in mammals – see e.g., Newey, Thirgood, & Hudson, 2004; Pioz et al., 2008; Stoehr & Kokko, 2006; Wasser & Barash, 1984) and their reproductive variance is much lower than that of males (Bateman, 1948; Trivers, 1972; see also susceptible male hypothesis: Nunn et al., 2009; Rolff, 2003; Stearns, 1992; Stoehr & Kokko, 2006).

With these insights in mind, the relationship between stressful early life environments and multiple aspects of immune function were investigated in men and women. The current research was designed to extend previous work linking early life environments to adult immune function to address three key questions: (a) What are the critical features of stressful early life environments that drive the association between childhood stress and adult immune function?, (b) How do the effects of these environmental stressors manifest themselves across qualitatively distinct aspects of immunological function?, and (c) Are these patterns sex-differentiated?

To address question *a*, the relative impact of three related, but distinct, sources of childhood stress on immune function were examined, including: (1) resource availability (i.e., SES: Griskevicius et al., 2011), (2) unpredictability (Mittal et al., 2015), and (3) adverse childhood experiences (ACEs: WHO, 2009). Isolating the unique impact that each of these environmental correlates of stress has on adult immune function will yield needed insights into the types of environmental interventions that are likely to have a lasting positive impact on adult health outcomes.

Regarding question *b*, much of the previous work linking childhood environments to adult immune function has focused primarily on inflammatory processes (e.g., Levine, Cole & Weir, 2015; Miller & Chen, 2010; Miller et al., 2019). In the current research, data were collected on a variety immunological endpoints, many of which were measured at multiple time points or at multiple effector-to-target (E:T) ratios. These measures included 1) proliferation of peripheral blood mononuclear cells (PBMCs) in response to, and in the absence of, stimulation by lipopolysaccharide (LPS), phytohemagglutinin (PHA), and polyinosinic:polycytidylic acid (poly [I:C]) measured at 24, 48, and 72 hrs post-plating; 2) natural killer (NK) cell lysis of target tumor cells at 100:1, 50:1, 25:1, and 12.5:1 E:T ratios; 3) phagocytosis of opsonized *Escherichia coli* (*E. coli*) bioparticles; and 4) PBMC release of the pro-inflammatory cytokines interleukin-1β (IL-1β), interleukin-6 (IL-6), and tumor-necrosis factor-α (TNF-α) in response to LPS stimulation, as well as in the absence of stimulation (i.e., spontaneous release), at 2, 24, 48, and 72 hrs post-plating. Additionally, plasma levels of total testosterone were also assayed to test whether diminished immune function co-occurs with increased investment in mating effort.

Lastly, to address question *c*, in line with the National Institute of Health’s recent policy on accounting for sex as a biological variable (see NIH, 2020), a number of steps were taken during study design, data collection, and data analysis to allow for targeted tests of sex differences in the impact of early life stress on immune function. These included recruiting equal numbers of men and women (with women all being naturally cycling and in the early follicular phase of their menstrual cycle), testing interactions between each predictor and sex in all analyses (i.e., rather than merely controlling for sex), and clearly presenting sex-differentiated results (see Table 2).

**Table 1.** Descriptive Statistics for Predictors and Immunological Measures (N = 159)

| **Variable** | ***M (SD)*** |
| --- | --- |
| Childhood SES | 4.55 (1.48) |
| Childhood Unpredictability | 2.37 (1.23) |
| Adverse Childhood Experiences | 3.51 (2.89) |
| Phagocytosis of *E. coli* bioparticles (%) | 12.59 (5.89) |
| NK Cell Cytotoxicity (%) | 37.58 (20.87) |
| Unstimulated Proliferation | .67 (.17) |
| Proliferation in Response to LPS | 1.01 (.22) |
| Proliferation in Response to PHA | 1.08 (.24) |
| Proliferation in Response to Poly (I:C) | .80 (.17) |
| Unstimulated Release of IL-1β | 52.62 (207.37) |
| Unstimulated Release of IL-6 | 307.01 (1031.63) |
| Unstimulated Release of TNF-α | 135.15 (200.14) |
| LPS-stimulated Release of IL-1β | 1417.09 (1288.25) |
| LPS-stimulated Release of IL-6 | 4972.26 (2283.22) |
| LPS-stimulated Release of TNF-α | 1931.45 (1136.58) |

*Note.* Peripheral blood mononuclear cell (PBMC) proliferation values shown here as absorbance reading at 490 nm collapsed across time-points (24 hr, 48 hr, 72 hr) for each plating condition. Values for natural killer cell lysis of K-562 tumor cells (i.e., NK cell cytotoxicity) shown here collapsed across effector:target cell ratios (100:1, 50:1, 25:1, 12.5:1). Levels of cytokines released from PBMCs *in vitro* shown here as pg/mL. SES = socioeconomic status; LPS = lipopolysaccharide, PHA = phytohemagglutinin, poly (I:C) = polyinosinic:polycytidylic acid, NK = natural killer, *E. coli* = *Escherichia coli*, IL-1β = interleukin-1 beta, IL-6 = interleukin-6, TNF-α = tumor necrosis factor-alpha.

**Table 2.** Summary of Sex-Specific Simple Slopes and Main Effects of Childhood Socioeconomic Status.

| Dependent Variable | Men |  |  | Women |  |  |
| --- | --- | --- | --- | --- | --- | --- |
|  | Coefficient (SE) |  |  | Coefficient (SE) |  |  |
|  | Primary Model | Covariates Model | Partial Model | Primary Model | Covariates Model | Partial Model |
| **Phagocytosis** | .31 (.12)** | .30 (.15)* | .29 (.13)* | -.09 (.12) | -.05 (.14) | -.17 (.14) |
| **Testosterone** | -.24 (.06)*** | -.22 (.07)** | -.25 (.07)*** | -.11 (.13) | -.21 (.14) | -.08 (.15) |
| **NK Cell Cytotoxicity** |  |  |  |  |  |  |
| Intercept | .19 (.14) | .09 (.18) | .20 (.11)† | -.17 (.12) | -.34 (.13)* | -.12 (.12) |
| Linear Slope | .39 (.10)*** | .42 (.18)* | .40 (.10)*** | -.18 (.12) | -.18 (.14) | -.37 (.13)** |
| Quadratic Slope | -.33 (.11)** | -.52 (.18)** | -.36 (.12)** | .11 (.13) | .11 (.18) | .32 (.13)* |
| *No significant interactions with sex below (main effects for sexes combined reported)* |  |  |  |  |  |  |
| **Proliferation - LPS** |  |  |  |  |  |  |
| Intercept | .19 (.09)* | .10 (.10) | .22 (.10)* |  |  |  |
| Slope | .08 (.13) | -.22 (.18) | .12 (.15) |  |  |  |
| **Proliferation - PHA** |  |  |  |  |  |  |
| Intercept | .15 (.06)* | .12 (.08) | .17 (.07)** |  |  |  |
| Slope | -.08 (.07) | -.19 (.10) | -.09 (.08) |  |  |  |
| **Proliferation - Poly (I:C)** |  |  |  |  |  |  |
| Intercept | .21 (.09)* | .10 (.10) | .22 (.10)* |  |  |  |
| Slope | -.01 (.01) | -.41 (.33) | -.02 (.02) |  |  |  |
| **PBMC Cytokine Release** |  |  |  |  |  |  |
| Unstimulated | -.22 (.07)** | -.22 (.09)* | -.33 (.09)*** |  |  |  |
| Stimulated | .04 (.05) | .03 (.04) | .03 (.05) |  |  |  |

*Note.* Standardized beta coefficients and standard errors (SEs) shown for main effects (no significant interaction found; bottom panel) and simple slopes within each sex for interactions found between childhood socioeconomic status and sex (top panel). NK = natural killer, LPS = lipopolysaccharide, PHA = phytohaemagglutinin, poly (I:C) = polyinosinic:polycytidylic acid, PBMC = peripheral blood mononuclear cells. † $p < .07$ , \* $p < .05$ , \*\* $p < .01$ , \*\*\* $p < .001$ .

It was hypothesized that growing up in adverse environments would have an especially negative impact on the immune function of men, with less pronounced effects found in women. Moreover, consistent with insights from the evolutionary sciences (e.g., the susceptible male hypothesis: Nunn et al., 2009; Rolff, 2003; Stearns, 1992; Stoehr & Kokko, 2006), it was predicted that this effect would co-occur with upregulation of testosterone, a key hormone driving mating effort (Alvergne, Faurie, & Raymond, 2009; Bancroft, 2005; Shirazi et al., 2019).

## Data Analysis Plan

See supplemental materials for additional details about data analysis. All models were estimated using MPlus statistical software (MPlus Version 8; Muthén & Muthén, 2012). All significance tests were two-tailed. Models were tested iteratively. First, relationships between childhood environmental factors (i.e., childhood SES, unpredictability, and adverse experiences), sex, and each dependent measure were tested in separate models for each environmental predictor. The results of these models are reported below as the primary analyses (see Results). Next, each model containing significant predictors was tested a second time while controlling for standard covariates, including adult SES (see Method for full list). Lastly, a model for each dependent measure was tested that examined the effects of all childhood environmental factors (and their interactions with sex) on the outcome simultaneously (i.e., to obtain partial effects). These latter two models were tested to examine whether significant effects of childhood factors on the immunological outcomes were robust to controlling for the other distinct, but related environmental constructs, as well as potential covariates; results of these models can be found in the supplemental materials. Information about whether the pattern or significance of the results of the primary analyses differed from these follow-up analyses are reported in the main text and Table 2.

For each model, the dependent measure(s) were regressed on the childhood environmental factor, sex, and the interaction between sex and the childhood environment factor. Per convention, if no significant interaction was found, main effects were reported without inclusion of the interaction term. Where significant interactions were found, direct effects of the childhood factor(s) on the dependent immunological measures within each sex (i.e., simple slopes) were examined. Sex differences at high and low levels (i.e., one standard deviation above and below the mean) of childhood factors involved in a significant interaction with sex in the initial models (i.e., regions of significance) were also probed. Given that the primary objective of the current study was to test whether the impact of childhood environmental factors on immune function differed by sex, only the simple slopes within each sex are reported in the main text. Regions of significance analysis can be found in the supplemental materials and Figures 1–2.

**Figure 1.**
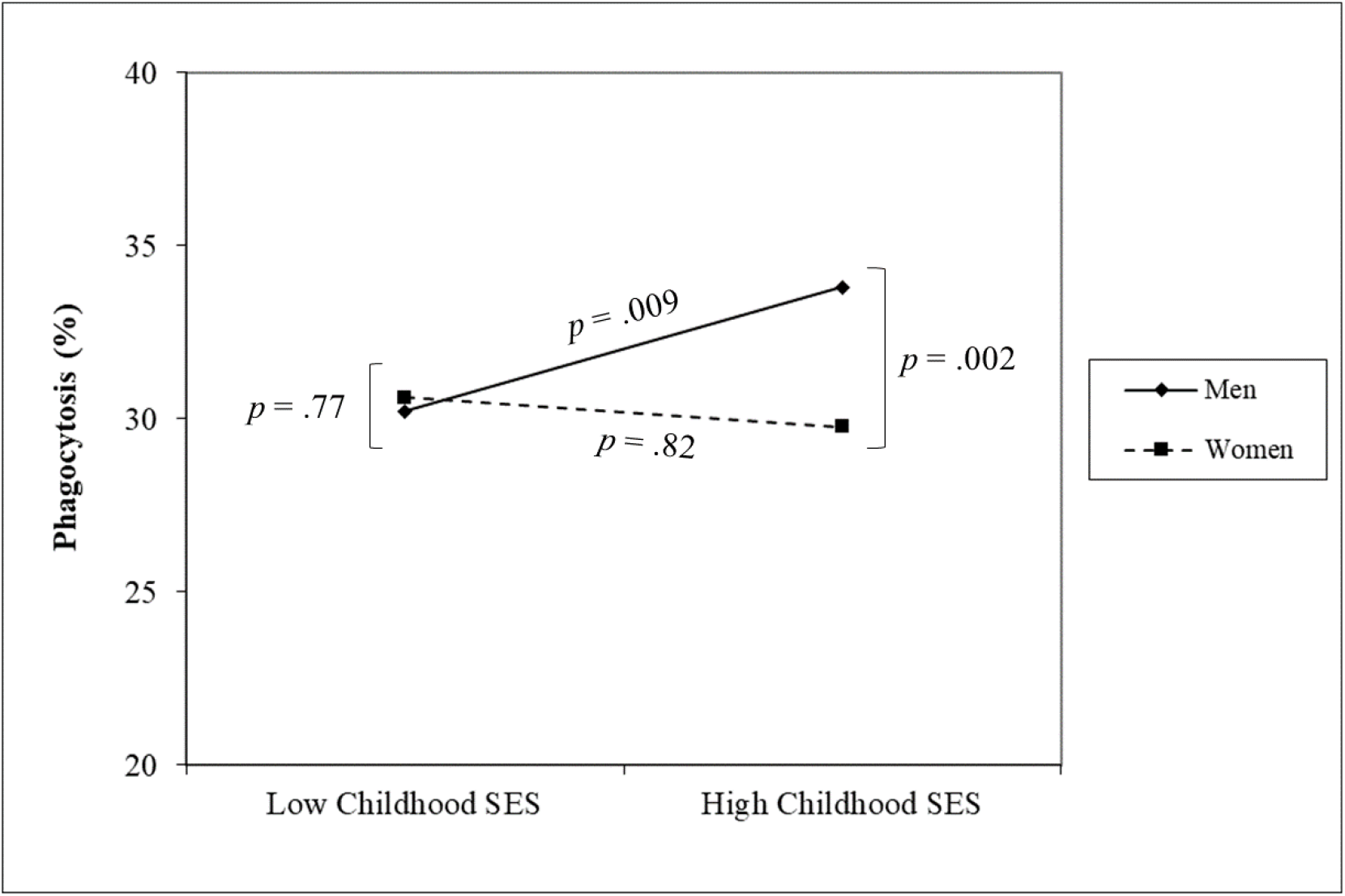
Interaction between childhood socioeconomic status (SES) and sex on phagocytosis of *Escherichia coli* bioparticles by peripheral blood mononuclear cells. High and low SES refer to one standard deviation above and below the mean of this variable, respectively.

**Figure 2.**
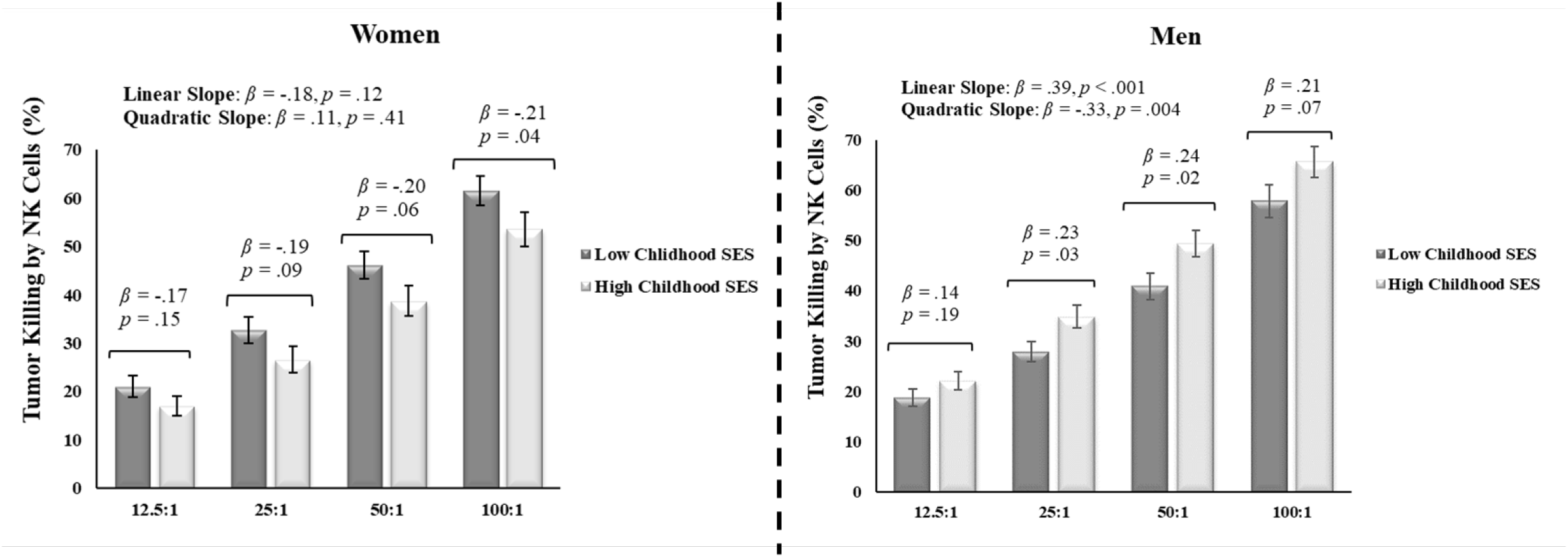
Interaction between childhood socioeconomic status (SES) and sex on natural killer (NK) cell killing of tumor cells (i.e., cytotoxicity) across effector:target cell ratios (12.5:1, 25:1, 50:1, and 100:1). Error bars reflect standard errors. High and low childhood SES refer to one standard deviation above and below the mean of this variable, respectively.

Given that the proliferation (time collected), NK cell cytotoxicity (E:T ratios), and cytokine release (time collected) data all contained nested structures, complex multilevel modeling (cytokine release data) and latent curve models (i.e., using structural equation modeling framework; proliferation and NK cell cytotoxicity data) were used to analyze these data. First, unconditional models for each of the dependent measures (i.e., without predictors included) were tested to assess model fit to linear change across repeated measures (see Table S1 for model fit indices). Model fit to linear change was acceptable for the proliferation data, so predictors were added to this model. For the NK cell cytotoxicity, model fit was poor to linear change, but improved to acceptable fit after the addition of a quadratic change slope. For the cytokine release data, given the large amount of missing data and non-linear change from the first to last time point that did not fit well to linear or quadratic times scores (see Figures S1–S2 in supplemental materials for demonstration), spontaneous cytokine release was modeled as a latent factor comprised of release of the three cytokines across time-points in the absence of stimulation (standardized factor loadings – IL-6: .97; IL-1β: .94; TNF-α: .90), and stimulated cytokine release was modeled as a latent factor comprised of release of the three cytokines in response to LPS stimulation across time points (standardized factor loadings – IL-6: .94; IL-1β: .92; TNF-α: .75). The phagocytosis and total testosterone data did not contain nested structures, and were thus analyzed together in a single-level, multivariate model.

## Results

Results of all models revealed good model fit (see Table S1 in supplemental materials for model fit statistics). Descriptive statistics are shown in Table 1. Results revealed that neither childhood unpredictability, nor cumulative exposure to adverse childhood experiences were related to any of the immunological measures, testosterone levels, nor did they interact with participant sex to impact any of these outcomes (all *p*s > .10; see supplemental materials for these results).^1^ Additionally, no factors or interactions emerged as significant predictors of unstimulated PBMC proliferation or stimulated *in vitro* cytokine release.

As predicted based on research indicating greater male susceptibility to adverse health effects of early life stress, results revealed significant interactions between childhood SES and sex in predicting NK cell cytotoxicity (intercept: *β* = -.54, *SE* = .28, *t* = -1.97, *p* = .049; linear slope: *β* = -.97, *SE* = .26, *t* = -3.68, *p* < .001; quadratic slope: *β* = .72, *SE* = .30, *t* = 2.38, *p* = .018), phagocytosis of *E. coli* bioparticles, *β* = -.64, *SE* = .27, *t* = -2.35, *p* = .02, and testosterone levels, *β* = .38, *SE* = .14, *t* = 2.73, *p* = .006.

Unpacking these interactions (see Figures 1–2 and Table 2) revealed that, for men, higher childhood SES predicted greater phagocytosis of *E. coli* bioparticles, *β* = .31, *SE* = .12, *t* = 2.62, *p* = .009, greater NK cell cytotoxicity, particularly at the two intermediate E:T ratios (intercept: *β* = .14, *SE* = .11, *t* = 1.31, *p* = .19; linear slope: *β* = .39, *SE* = .10, *t* = 4.03, *p* = .001; quadratic slope: *β* = -.33, *SE* = .11, *t* = -2.90, *p* = .004), and lower levels of testosterone, *β* = -.24, *SE* = .06, *t* = -3.79, *p* < .001. For women, childhood SES did not significantly predict phagocytosis, *β* = -.09, *SE* = .12, *t* = -.22, *p* = .82, NK cell cytotoxicity (intercept: *β* = -.17, *SE* = .12, *t* = -1.45, *p* = .15; linear slope: *β* = -.18, *SE* = .12, *t* = -1.57, *p* = .12; quadratic slope: *β* = .11, *SE* = .13, *t* = .83, *p* = .41), or testosterone levels, *β* = -.11, *SE* = .13, *t* = -.87, *p* = .38. Notably, although only reaching statistical significance at the highest E:T ratio (see Figure 2), the relationship between childhood SES and NK cell cytotoxicity in women was in the opposite direction as was observed in men. Specifically, in women, higher childhood SES was related to reduced NK cell cytotoxicity (see supplemental materials for a discussion of this pattern). The pattern and significance of these results remained largely unchanged when controlling for covariates (including adult SES) or by controlling for the effects of other childhood environmental variables (see Table 2 and supplemental materials).

In addition to these sex-differentiated results, significant main effects of childhood SES (i.e., across sexes) were found predicting the intercept of PBMC proliferation (i.e., at the first time-point of 24 hrs) in response to stimulation by LPS, *β* = .19, *SE* = .09, *t* = 2.17, *p* = .03, PHA, *β* = .15, *SE* = .06, *t* = 2.32, *p* = .02, and poly (I:C), *β* = .21, *SE* = .09, *t* = 2.44, *p* = .02, but not the slopes of proliferation over time in these conditions (*p*s > .27). These effects were no longer significant after controlling for covariates (*p*s > .12; see supplemental materials), and should thus be interpreted with caution. Lastly, there was a main effect of childhood SES on the latent factor of spontaneous *in vitro* proinflammatory cytokine release by PBMCs (i.e., cells plated in media only), *β* = -.22, *SE* = .07, *t* = -3.12, *p* = .002, but not stimulated cytokine release, *β* = .04, *SE* = .05, *t* = .86, *p* = .39. Specifically, higher childhood SES predicted a diminished tendency to release proinflammatory cytokines in the absence of overt immunological stimulation. These results remained significant when controlling for both covariates and the effects of the other childhood environmental factors.

## Discussion

See supplemental materials for an expanded discussion. Much research indicates that early life experiences have a lasting impact on adult health (Delpierre et al., 2018; Levine et al., 2015; Miller & Chen, 2010; Miller et al., 2019; Taylor, 2010), especially for men (Aiken & Ozanne, 2013; Conti et al., 2015; Del Giudice et al., 2018; Campbell et al., 2014; Garcia et al., 2016; Geary, 2016; Stinson, 1985). The current research sought to extend this work by addressing three key questions that have been heretofore unexplored in the context of a single investigation: (a) What are the critical features of early life stress that drive the association between childhood stress and adult health outcomes?, (b) How do these factors manifest themselves across qualitatively distinct aspects of immunological function?, and (c) Are these patterns sex-differentiated?

Results revealed that men exposed to lower childhood SES exhibited a diminished immune response across each measure of immunological function, as well as increased spontaneous pro-inflammatory cytokine release by PBMCs, compared to men with a higher childhood SES. Conversely, with the exception of the effects of childhood SES on stimulated PBMC proliferation and spontaneous cytokine release (both effects not sex-differentiated), the measured childhood environmental factors did not significantly predict immunological function in women. Further, with the exception of the relationship between childhood SES and stimulated PBMC proliferation, significant effects were not attenuated when controlling for adult SES (or other covariates; see supplemental materials for these results), suggesting that these differences are primarily the result of developmental processes rather than current resource availability.

The present results are consistent with a growing body of research demonstrating a negative impact of early life adversity on adult health (Delpierre et al., 2018; Levine et al., 2015; Miller & Chen, 2010; Miller et al., 2019; Taylor, 2010), as well as research demonstrating sex differences in key immunological outcomes (Nunn, 2009; Rolff, 2003; Stoehr & Kokko, 2006). Further, consistent with evolutionary biological models that predict tradeoffs between immune function and mating effort (French, DeNardo, & Moore, 2007; McKean & Nunney, 2011; Norris & Evans, 2000; Stoehr & Kokko, 2006), results revealed that, in men, reduced investment in immunological functioning co-occurs with increased investment in testosterone levels. The results of the current research suggest that men who grow up in conditions of low SES may be more vulnerable to poor health outcomes later in life, due to diminished investment in immune function relative to investment in mating effort. Moreover, the current results suggest that these differences might emerge in response to differential resource access, per se, as there was little evidence of a negative impact of childhood unpredictability or other adverse experiences (e.g., abuse, neglect) on immune function in either sex^1^.

There are limitations of the current research that should be considered. First, although several components of immune function were measured in the current study, together they capture only a narrow range of the tremendous complexity that comprises immune function. Accordingly, future research might find that, in women, early life experiences play an important role in influencing other aspects of immune function, such as adaptive immunity. Further, the current research did not experimentally examine the proximate biological mechanisms driving the observed relationships between early life stress and adult immune function. One possibility is that glucocorticoids and other adrenal hormones (e.g., dehydroepiandrosterone [DHEA]) play an important role in this context, as they have immunomodulatory properties (Buford & Willoughby, 2008) and levels of these hormones are impacted by early life stress (Van Voorhees et al., 2014). It is also important to consider that the majority of the participants in the current study were healthy college students whose exposure to early life stress may have been lower than that of the general population. Moreover, all of measures of early life stress used in this study were retrospective. Thus, a key direction for future research will be to examine the effects of childhood SES and multiple types of early life adversity on adult immune function in a more diverse sample, perhaps using a longitudinal design. On that note, future research may also benefit from utilizing longitudinal designs to track sex differences in the impact of early life conditions on immune function across multiple stages of development.

The current research provides important evidence of sex differences in the impact of early life SES on multiple facets of immune function. Together, these findings suggest that, for men in particular, early life experiences become immunologically embedded in ways that may have a lasting impact on health. These results have important implications for interventions targeted at reducing the negative impact of early life disadvantage on health, and lay the groundwork for future research examining sex differences in relationships between childhood environments and immune function across the lifespan.

## Materials and Methods

### Participants

A total of 159 individuals from Texas Christian University or the surrounding community, participated in exchange for $25 or experimental research credit (80 men, 79 women; *M*_age_ = 20.17 years, *SD* = 2.75). Restriction criteria were: 1) participants must have no history of chronic medical problems, including mental illness, 2) must be of healthy weight (i.e., body mass index [BMI] < 30), 3) must be free from acute illness for at least two weeks prior to participation, 4) women must not be on hormonal contraceptives, 5) must be willing to abstain from steroidal and non-steroidal anti-inflammatory medications, exercise, and alcohol use for at least two days before participating, and 6) must fast the morning of the session. Additional characteristics of the sample are published elsewhere (42). Women participated 4–7 days after the start of their most recent menstrual cycle (i.e., early follicular phase). In exchange for participation, participants were given the choice of partial course credit or a $50 gift card.

### Procedure

All participants provided informed consent prior to participation, and the study was approved as compliant with ethical standards by the Texas Christian University Institutional Review Board (Approval #: 1411-117-1606).

On the day of their sessions, participants arrived at the laboratory at 7:30 AM, provided informed consent, and answered study compliance questions (e.g., whether or not they fasted, abstained from alcohol use, etc.). Next, for the purpose of a larger study, participants completed a series of behavioral tasks and questionnaires. Finally, participants were led into an adjoining room, in which 85mL of blood was collected by venipuncture into heparinized (or EDTA-containing) Vacutainer® tubes (Becton-Dickinson, Franklin Lakes, NJ).

Peripheral blood mononuclear cells (PBMCs) were immediately isolated from whole blood using density gradient centrifugation in Ficoll® Paque Plus (Sigma-Aldrich, St. Louis, MO [GE Healthcare Life Sciences]). PBMCs were used in four functional immunoassays: 1) natural killer (NK) cell lysis of target tumor cells, 2) PBMC phagocytosis of fluorescent dye-labelled *Escherichia coli (E. coli)* bioparticles, 3) PBMC proliferation in response to, and in the absence of, mitogen / toll-like receptor stimulation, and 4) PBMC release of proinflammatory cytokines in response to, and in the absence of, mitogen stimulation. For each assay, PBMCs were brought to the plating density appropriate for the assay in RPMI-1640 supplemented with 10% heat-inactivated fetal bovine serum (FBS), 2mM L-glutamine, 1mM sodium pyruvate, 100 U of penicillin/mL, 100µg of streptomycin/mL, and 0.25µg of amphotericin B/mL (Caisson Labs, Logan, UT). In addition, plasma was collected at the time of blood processing and frozen at -80°C. Additional information about each assay follows, below.

## Materials

### Childhood environmental factors

First, participants’ childhood socioeconomic status (SES) was assessed using a previously validated scale (Griskevicius et al., 2011; see also Hill et al., 2016). This measure was chosen to capture the extent to which participants felt they did or did not have access to resources during their childhood. Participants indicated their childhood SES by responding to three statements about their life before age 12 (e.g., “My family usually had enough money for things when I was growing up”) using a 7-point Likert scale (1: *Strongly disagree*, 7: *Strongly agree*). The items together yielded acceptable reliability (α = .81) and were formed into a mean composite, with a higher score indicating greater resource availability / a higher SES during childhood (*M* = 4.55, *SD* = 1.48).

Next, childhood unpredictability was measured with a scale used in previous research (Mittal et al., 2015), consisting of three statements about chaos and uncertainty within the home (e.g., “Things were often chaotic in my house”). Participants reported agreement with how well each statement described their lives before age 12 using a 7-point scale (1: *Strongly disagree*, 7: *Strongly agree*). Together, the items yielded acceptable reliability (α = .74) and were formed into a mean composite with a higher score indicating greater experience with unpredictability during childhood (*M* = 2.37, *SD* = 1.23).

Finally, participants’ experience with other types of adversity during childhood were also measured using the well-validated Adverse Childhood Experience International Questionnaire (ACE-IQ), developed by research teams from the World Health Organization (WHO, 2009). This questionnaire assesses experience with adversity during childhood across five domains: abuse, family dysfunction, and peer, community, and collective violence/war. Given the nature of the present sample sample, which was primarily composed of college students, a truncated version of the survey was administered, removing the collective violence, sexual abuse, and secondary household abuse questions, and leaving a total of 20 items. Continuous items were recoded as binary, per convention, and all items were then summed into a composite measure of cumulative exposure to adverse experiences during childhood (*M* = 3.51, *SD* = 2.89).

### NK cell cytotoxicity assay

NK cell cytotoxicity was measured using the classic ^51^Cr-release assay. K-562 target tumor cells (ATCC^®^ CCL-24^TM^, Manassas, VA) were grown inside T-25 flasks (ThermoFisher Scientific, Waltham, MA) in RPMI-1640 growth medium supplemented with 10% FBS, 2mM L-glutamine, 1mM sodium pyruvate, 100 U of penicillin/mL, 100µg of streptomycin/mL, and 0.25µg of amphotericin B/mL (Caisson Labs, Logan, UT) and incubated at 37°C, 5% CO_2_, and 100% humidity.

Target tumor cells were labeled with 1 µCi ^51^Cr (PerkinElmer, Waltham, MA) for one hour before plating them at a density of 1 x 10^4^ cells/well in a 200µL final volume into Corning^®^ V-bottom plates (Corning, Corning, NY). PBMCs were plated with target tumor cells, in triplicate, at the following effector:target (E:T) cell ratios: 100:1 (1 x 10^6^ PBMCs/well), 50:1 (5 x 10^5^ PBMCs/well), 25:1 (2.5 x 10^5^ PBMCs/well), and 12.5:1 (1.25 x 10^5^ PBMCs/well). Spontaneous/background lysis controls (i.e., target cells plated in media only, with no PBMCs) and maximal lysis controls (i.e., target cells plated in 1% Triton X-100 [Sigma-Aldrich, St. Louis, MO]) were plated in sextuplicate. Plates were incubated for four hours at 37°C, 5% CO_2_, and 100% humidity.

After brief centrifugation of the V-bottom plate, supernatants were collected into glass scintillation vials and quantified ^51^Cr release on a CAPRAC®-t gamma counter (Capintec, Inc., Ramsey, NJ). Percent maximal lysis of target tumor cells by participant NK cells was calculated at each E:T ratio by dividing ^51^Cr release by maximal release after subtracting spontaneous release from both values, and multiplying by 100.

### Phagocytosis assay

The phagocytic capability of participants’ PBMCs was assessed using fluorescent pHrodo™ Green *E. coli* BioParticles™ (ThermoFisher Scientific, Waltham, MA) that were opsonized using 1 mg/mL of manufacturer-provided opsonization buffer. PBMCs were plated in triplicate with *E. coli* bioparticles into BrandTech® black, flat-bottom microplates (BrandTech Scientific, Essex, CT) at a density of 5 x 10^5^ cells/well in a 200µL final volume. Negative controls (bioparticles plated with media only in 200µL volume) and positive controls (bioparticles plated with pH 4.5 Intracellular Calibration Buffer [ThermoFisher Scientific, Waltham, MA]) were plated in triplicate. Plates were incubated for two hours at 37°C, 5% CO_2_, and 100% humidity before being read on a fluorescence plate reader (BMG LabTech FLUOstar™ Omega, Cary, NC) at FITC dye settings of 490nm excitation/ 520nm emission. Percent maximal fluorescence was computed by dividing experimental fluorescence by maximal fluorescence (i.e., positive control), after subtracting fluorescence in the negative controls from both, and multiplying by 100.

### PBMC proliferation assay

PBMCs were isolated as described above, and plated into Falcon^®^ 96-well tissue culture plates (Corning, Corning, NY). PBMCs were plated in triplicate at a final density of 2.5 x 10^5^ cells/well in a 200µL final volume for four conditions: with media only (i.e., unstimulated proliferation), 2) with 1µg/mL of lipopolysaccharide (LPS) obtained from *E. coli* (serotype 026:B6, Sigma-Aldrich, St. Louis, MO), 3) with 5µg/mL of phytohemagglutinin (PHA; Sigma-Aldrich, St. Louis, MO), and 4) with 50µg/mL of polyinosinic:polycytidylic acid (poly [I:C]; high molecular weight; InvivoGen, San Diego, CA). Plates were incubated at 37°C, 5% CO_2_, and 100% humidity.

Approximate cell density was measured at three time-points: 24, 48, and 72 hrs post-plating using the CellTiter 96^®^ AQueous Non-Radioactive Cell Proliferation Assay (Promega, Madison, WI). Each plate was read on a plate reader (BMG LabTech FLUOstar™ Omega, Cary, NC) at 490 nm.

### Proinflammatory cytokine release assay

PBMC release of proinflammatory cytokines was measured *in vitro* both in response to LPS stimulation, as well as in the absence of stimulation. After isolation, PBMCs were plated in triplicate at a density of 2.5 x 10^5^ cells/well, in a 200µL final volume. PBMCs were plated both in media only (unstimulated condition), as well as with 1µg/mL of LPS, obtained from *E. coli* (serotype 026:B6, Sigma-Aldrich, St. Louis, MO), and were incubated for up to three days at 37°C, 5% CO_2_, and 100% humidity. Cell culture supernatants were collected at 2, 24, 48, and 72 hrs post-plating, and then stored them at -80°C until assays were conducted.

Cell culture supernatants were later thawed and assayed in duplicate for levels of a trio of proinflammatory cytokines: interleukin-1beta (IL-1β), interleukin-6 (IL-6), and tumor necrosis factor-alpha (TNF-α) using a MILLIPLEX® MAP Human Cytokine Panel magnetic bead kit (EMD Millipore Corporation, Billerica, MA), and read using a Luminex MAGPIX® fluorescent detection system (Luminex, Austin, TX) and xPONENT® software (Version 4.2; build: 1324; Luminex, Austin, TX). Intra-assay coefficients of variation (CVs) were 8.20% (IL-6), 6.97% (IL-1β), and 5.98% (TNF-α). Inter-assay coefficients of variation (CVs) were 17.27% (IL-6), 10.53% (IL-1β), and 11.62% (TNF-α).

Due to a freezer failure on November 5, 2016, cell culture supernatant samples were compromised for 32 participants (see also Gassen et al., 2019 for additional information). No compromised samples were assayed, and thus data from these samples were not included in any analysis. All other biological samples from these participants (including plasma used for the total testosterone assay) were stored elsewhere and were thus unaffected by the freezer failure.

### Total testosterone assay

Plasma samples were and assayed in duplicate for levels of total testosterone using commercially-available enzyme-linked immunosorbent assay kits (Abcam, Cambridge, UK). Plates were read on a plate reader (BMG LabTech FLUOstar™ Omega, Cary, NC) at a wavelength of 450 nm. The intra-assay CV was 2.48% and the inter-assay CV was 13.45%.

### Covariates and alternative explanations

Several variables that may influence relationships between early life conditions, sex, and immune function were measured. These included race, age, exercise, sleep, BMI, stress, loneliness, recent illness, season, and adult SES. See supplemental materials for more information.

## Data Availability

Data will be available on the Open Science Framework at the time of publication (DOI 10.17605/OSF.IO/DXPZU).

## Footnotes

^1^While neither childhood unpredictability, nor adverse childhood experiences, significantly predicted any immunological outcome in the primary models or models controlling for covariates, results of the partial effects model revealed significant interaction between each of these variables and sex in predicting PHA-stimulated proliferation. Further, there was a main effect of childhood unpredictability on spontaneous cytokine release by PBMCs. These results can be found in the supplemental materials.

