## Supplemental materials for "Sex Differences in the Impact of Childhood Socioeconomic Status on Immune Function"

**This file includes:**

Supplementary Method and Results

Additional Discussion

Supplementary References

Table S1

Figure S1

**Supplementary Method and Results**

**Covariates**

Exercise was measured with the question, “How many hours of exercise do you do in a typical week?”, sleep quality was measured with the question, “How many hours of sleep do you get on a typical night?”, stress was measured using the Perceived Stress Scale (1), loneliness was measured using the revised UCLA Loneliness Scale (2), recent illness was measured with three items: (a) “I am feeling sick today”, (b) “I have felt sick within the past week”, and (c) “When was the last time you had a cold, flu, or other illness?”, season was measured using day length at the time of participation in Fort Worth, TX, and adult SES was measured using the McArthur Subjective Social Status Ladder (3).

**Data Analysis Plan**

Outlying values across the biological measures were excluded pairwise from data analysis (i.e., > 3 standard deviations above group mean; less than 2.8% values for any given measure: 3 phagocytosis values, 2 testosterone values, 14 proliferation values [across media-only, LPS, PHA, and poly [I:C] plating conditions and all time-points]; 4–5). The cytokine release data were positively skewed at each time point, and were thus log-transformed for all analyses, as this transformation corrected the skew.

Because the phagocytosis and testosterone data lacked a nested structure, these outcomes were simultaneously regressed on the predictors in a single-level model. The remaining data all contained nested structures. For the proliferation data, time-points (24, 48, and 72 hrs post-plating) were nested within each plating condition (i.e., media-only, LPS, PHA, and poly [I:C]), which were themselves nested within participants. For the NK cell cytotoxicity data, E:T ratios (100:1, 50:1, 25:1, and 12.5:1) were nested within each participant. For the stimulated cytokine release data, time-points (2, 24, 48, and 72 hrs post-plating) were nested within each plating condition (i.e., unstimulated or stimulated), which were themselves nested within participants. Thus, the multilevel and structural equation modeling techniques described in the main text were used to analyze these data.

Model fit was assessed using four fit indices: *χ*^2^ test of model fit, the comparative fit index (CFI), the root mean square error of approximation (RMSEA), and the standardized root mean square residual (SRMR). Adequate model fit was indicated by a non-significant *χ*^2^ value (*p* > .05), a CFI value > .90, an RMSEA value < .08, and an SRMR statistic < .08. Given that the *χ*^2^ value is often inflated with relatively large sample sizes, models with a significant *χ*^2^ value were still considered as having good fit as long as all other fit indices were within the specified range.

**Results of Models Testing Effects of Childhood Environmental Factors Separately**

**PBMC Proliferation**

For the PBMC proliferation models, time was centered at the first measured time-point (i.e., 24 hrs), such that the random intercept represented proliferation at 24 hrs. The linear slope term represented change in proliferation from 24 hrs to 72 hrs. Results revealed no significant interactions between any childhood environment variable and sex in predicting the intercept or slope of spontaneous PBMC proliferation (i.e., cells plated in media only; *ps* > .30). The main effect of sex on the intercept of spontaneous proliferation approached significance, *β* = .18, *SE* = .09, *t* = 1.94, *p* = .053, with women exhibiting a greater value than men. However, this effect no longer approached significance after controlling for covariates (see ‘Covariates’ section; *p* = .88). Sex did not significantly predict the slope of proliferation over time (*p* = .36). There were no significant main effects of any childhood measure on the intercept or slope of spontaneous proliferation (*p*s > .14). Final models explained between 3.2-3.4% of the variance in the intercept and 1.5–3.8% of the variance in the slope, depending on the childhood measure included in the model.

For proliferation in response to LPS stimulation, results revealed no significant interactions between any childhood environment variable and sex in predicting the intercept or slope (*p*s > .15). There were no significant main effects of sex (*p*s > .48). The main effect of childhood SES on the intercept of proliferation in response to LPS stimulation was significant, with higher childhood SES predicting greater proliferation measured at 24 hrs, *β* = .19, *SE* = .09, *t* = 2.17, *p* = .03. However, while the pattern of the relationship remained the same, this effect was no longer significant after controlling for covariates (*p* = .29). Childhood SES did not predict the slope of proliferation in response to LPS over time (*p* = .51). Neither childhood unpredictability nor adverse childhood experiences predicted the intercept or slope of proliferation in response to LPS over time (*p*s > .39). Final models explained between 0.5-3.8% of the variance in the intercept and 1.1–1.9% of the variance in the slope, depending on the childhood measure included in the model.

For proliferation in response to PHA stimulation, results revealed no significant interactions between any childhood environment variable and sex in predicting the intercept or slope (*p*s > .26). Sex did not significantly predict the intercept or slope (*p*s > .11). The main effect of childhood SES on the intercept of proliferation in response to PHA stimulation was significant, *β* = .15, *SE* = .06, *t* = 2.32, *p* = .02, with higher childhood SES predicting greater proliferation measured at 24 hrs. However, while the pattern of the relationship remained the same, this effect was no longer significant after controlling for covariates (*p* = .12). Childhood SES did not significantly predict the slope of proliferation over time (*p* = .27). Neither childhood unpredictability nor adverse childhood experiences predicted the intercept or slope of proliferation in response to PHA over time (*p*s > .30). ). Final models explained between 0.4–2.4% of the variance in the intercept and 1.2–1.8% of the variance in the slope, depending on the childhood measure included in the model.

For proliferation in response to poly (I:C) stimulation, there were no significant interactions between any childhood environment variable and sex in predicting the intercept or slope (*p*s > .24). Sex did not significantly predict the intercept or slope (*p*s > .68). The main effect of childhood SES on the intercept of proliferation in response to poly (I:C) stimulation was significant, *β* = .21, *SE* = .09, *t* = 2.44, *p* = .02, with higher childhood SES predicting greater proliferation. However, while the pattern remained the same, this effect was no longer significant after controlling for covariates (*p* = .30). Childhood SES did not significantly predict the slope of proliferation over time (*p* = .73). Neither childhood unpredictability nor adverse experiences predicted the intercept or slope of proliferation over time in this plating condition (*p*s > .36). Final models explained between 0.1–4.5% of the variance in the intercept and 0.1–1.0% of the variance in the slope, depending on the childhood measure included in the model.

**Phagocytosis**

For phagocytosis of *E. coli* bioparticles, the main effect of sex did not reach significance in any model (*p*s > .10). Neither the main effect of childhood unpredictability (*p* = .63), nor the main effect of adverse childhood experiences (*p* = .18) reach significance. The interactions between sex and these variables also did not reach significance (*p*s > .43). Results revealed a significant two-way interaction between sex and childhood SES, *β* = -.64, *SE* = .27, *t* = -2.35, *p* = .02. The model including childhood SES and its interaction with sex explained 7.5% of the variance in phagocytosis.

Specifically, in men, higher childhood SES predicted greater phagocytosis, *β* = .31, *SE* = .12, *t* = 2.62, *p* = .009. Childhood SES did not significantly predict phagocytosis in women, *β* = -.09, *SE* = .12, *t* = -.22, *p* = .82. At low childhood SES (1 standard deviation below the mean of this variable), men and women did not differ in phagocytosis (*p* = .77). However, at high childhood SES, women exhibited significantly reduced phagocytosis compared to men, *β* = -.34, *SE* = .11, *t* = -3.05, *p* = .002. The pattern and significance of these results did not change when covariates were controlled for. Specifically, the interaction between sex and childhood SES (*p* = .03), the effect of childhood SES on phagocytosis in men (*p* = .04), and the effect of sex on phagocytosis at high childhood SES (*p* = .001) all remained significant.

**Natural Killer Cytotoxicity**

For NK cell cytotoxicity, there were no significant main effects of either childhood unpredictability or adverse childhood experiences on the intercept, linear slope, or quadratic slope of tumor killing across E:T ratios (*p*s > .26). There were also no significant main effects of sex (*p*s > .15) or interactions between sex and these environmental variables (*p*s > .22) on any dependent measure. However, results revealed significant interactions between childhood SES and sex in predicting each the intercept, *β* = -.54, *SE* = .28, *t* = -1.97, *p* = .049, linear slope, *β* = -.97, *SE* = .26, *t* = -3.68, *p* < .001, and quadratic slope, *β* = .72, *SE* = .30, *t* = 2.38, *p* = .018. The model including childhood SES and its interaction with sex explained 3.0% of the variance in the intercept, 9.6% of the variance in the linear slope, and 6.0% of the variance in the quadratic slope.

Unpacking this interaction revealed that, in men, the effect of childhood SES on the intercept of NK cell cytotoxicity did not reach significance, *β* = .14, *SE* = .11, *t* = 1.31, *p* = .19. This suggests that childhood SES does not predict NK cell cytotoxicity at the lowest E:T ratio (i.e., 12.5:1). However, childhood SES significantly predicted both the linear slope, *β* = .39, *SE* = .10, *t* = 4.03, *p* = .001, and quadratic slope, *β* = -.33, *SE* = .11, *t* = -2.90, *p* = .004, of cytotoxicity across E:T ratios. Specifically, higher childhood SES predicted a greater increase in cytotoxicity as E:T ratios increased up to the 50:1 ratio (25:1 – *p* = .03; 50:1 – *p* = .02), which tapered off by the 100:1 ratio, *p* = .07. In women, childhood SES did not significantly predict the intercept, *β* = -.17, *SE* = .12, *t* = -1.45, *p* = .15, linear slope, *β* = -.18, *SE* = .12, *t* = -1.57, *p* = .12, or quadratic slope, *β* = .11, *SE* = .13, *t* = .83, *p* = .41. Notably, although non-significant, the relationship between childhood SES and NK cell cytotoxicity was in the opposite direction for women than it was for men.

At low childhood SES, sex did not predict the intercept, *β* = .10, *SE* = .12, *t* = .85, *p* = .40, or quadratic slope, *β* = -.17, *SE* = .13, *t* = -1.27, *p* = .21. However, sex did significantly predict the linear slope, *β* = .22, *SE* = .11, *t* = 2.00, *p* = .046, with women demonstrating a greater increase in NK cell cytotoxicity across E:T ratios than men at low levels of SES. At high childhood SES, men exhibited marginally greater NK cell cytotoxicity at the lowest E:T ratio (intercept), *β* = -.22, *SE* = .11, *t* = -1.94, *p* = .053. Sex also predicted both the linear, *β* = -.35, *SE* = .12, *t* = -2.95, *p* = .003, and quadratic slopes, *β* = .26, *SE* = .12, *t* = 2.17, *p* = .03, with men demonstrating greater NK cell cytotoxicity at all E:T ratios (25:1 – *p* = .01; 50:1 – *p* = .01; 100:1 – *p* = .02), with the effect being weaker at the highest E:T ratio.

The pattern and significance of these results were largely unchanged when covariates were controlled for. The interaction between childhood SES and sex did not significantly predict the intercept (*p* = .25), but continued to predict the linear slope (*p* = .04) and quadratic slope (*p* = .02). In men, childhood SES continued to significantly predict both the linear slope (*p* = .02) and quadratic slope (*p* = .003), but not the intercept (*p* = .61). In women, childhood SES did not predict either slope term (*p* > .19), but did significantly predict the intercept, *β* = -.23, *SE* = .09, *t* = -2.59, *p* = .01, with higher childhood SES predicted lower NK cell cytotoxicity at the 12.5:1 ratio. At low childhood SES, sex did not significantly predict any term (*p* > .09). As in the primary analysis, at high childhood SES, the relationship between sex and the intercept was marginally significant (*p* = .07). Sex significantly predicted the linear slope (*p* = .02), but not the quadratic slope (*p* = .10), indicating a greater increase in NK cell cytotoxicity as E:T ratios increased for men compared to women.

**PBMC Cytokine Release**

Results revealed that neither spontaneous, nor LPS-stimulated cytokine release were predicted by sex (*p*s > .28), adverse childhood experiences (*p*s > .73), or childhood unpredictability (*p*s > .07). There were also no significant interactions between sex and any childhood environmental measure (*p*s > .39). However, results revealed that higher childhood SES significantly predicted reduced spontaneous cytokine release (i.e., from cells plated in media only), *β* = -.22, *SE* = .07, *t* = -3.12, *p* = .002, accounting for 5.0% of the variance in this outcome. Childhood SES, however, did not significantly predict stimulated cytokine release, *β* = .04, *SE* = .05, *t* = .86, *p* = .39 (explaining 0.1% of variance in outcome). The pattern and significance of these results remained the same after controlling for covariates, with higher childhood SES continuing to significantly predict lower spontaneous cytokine release (*p* = .02), but not stimulated cytokine release (*p* = .50).

**Testosterone**

As expected, the main effect of sex was significant, such that men had higher testosterone levels than women, *β* = -1.22, *SE* = .13, *t* = -9.70, *p* < .001. Neither adverse childhood experiences (*p* = .21), nor childhood unpredictability (*p* = .11) significantly predicted testosterone levels. Further, the two-way interactions between sex and each of these variables did not reach statistical significance (*p*s > .29). However, results revealed a significance two-way interaction between sex and childhood SES, *β* = .38, *SE* = .14, *t* = 2.73, *p* = .006. The model including childhood SES and its interaction with sex explained 78.4% of the variance in testosterone levels.

Unpacking this interaction revealed that men had higher levels of testosterone than women both at high childhood SES, *β* = -.76, *SE* = .05, *t* = -15.03, *p* < .001, and low childhood SES, *β* = -.99, *SE* = .04, *t* = -22.24, *p* < .001. Further, in men, higher childhood SES predicted significantly lower testosterone levels, *β* = -.24, *SE* = .06, *t* = -3.79, *p* < .001. No significant relationship was found between childhood SES and testosterone levels in women, *β* = -.11, *SE* = .13, *t* = -.87, *p* = .38. The pattern and significance of these did not change when covariates were controlled for. Specifically, both the two-way interaction between sex and childhood SES (*p* = .03), as well as the effect of childhood SES on testosterone in men (*p* = .001), each remained statistically significant.

**Results of Testing Partial Effects of Childhood Environmental Variables**

As described in the main text, each of the models was re-tested with all childhood environmental variables (and their interactions with sex) included in the model simultaneously. This analysis was conducted to determine whether the pattern of the relationships between each childhood environmental variable, sex, and each outcome differed when the effects of the other environmental variables were controlled for. Given that the primary objective was to test whether the impact of childhood environmental factors on immunological outcomes differed by sex, significant interactions found in these analyses were only unpacked by examining simple slopes within each sex (for regions of significance, see previous set of analyses).

**PBMC Proliferation**

There were no significant main effects of any childhood variable on the intercept or slope of spontaneous PBMC proliferation (i.e., cells plated in media only) (*p*s > .10). Additionally, no interactions between any childhood environmental variable and sex significantly predicted either the intercept or slope (*p*s > .09).

For LPS-stimulated proliferation, only the main of effect of childhood SES on the intercept (but not slope, *p* = .45) was significant, *β* = .22, *SE* = .10, *t* = 2.29, *p* = .02, with higher SES predicting greater proliferation at 24 hrs. Neither childhood unpredictability (*p*s > .65), nor adverse childhood experiences (*p*s > .36) significantly predicted either the slope or the intercept of proliferation over time. Further, no interactions between childhood environmental variables and sex reached significance (*p*s > .08).

For PHA-stimulated proliferation, there was a significant main effect of childhood SES on the intercept, *β* = .17, *SE* = .97, *t* = 2.32, *p* = .02, but not slope (*p* = .35), with higher childhood SES predicting greater proliferation at 24 hrs. There were no significant interactions between childhood SES and sex (*p*s > .38). There were no significant interactions between childhood unpredictability or adverse childhood experiences and sex on the intercept of proliferation (*p*s > .12). However, sex significantly interacted with both unpredictability, *β* = .54, *SE* = .21, *t* = 2.54, *p* = .01, and adverse experiences, *β* = -.51, *SE* = .17, *t* = -3.09, *p* = .002, to predict the slope.

Results revealed that in men, childhood unpredictability did not significantly predict the slope of PHA-stimulated proliferation, *β* = -.37, *SE* = .26, *t* = -1.44, *p* = .15. However, in women, greater childhood unpredictability predicted a greater increase in proliferation over time, *β* = .65, *SE* = .27, *t* = 2.40, *p* = .02. With adverse childhood experiences, a greater number of adverse experiences reported predicted a greater increase in proliferation over time in men, *β* = .58, *SE* = .27, *t* = 2.17, *p* = .03, but a diminished increase in proliferation over time in women, *β* = -.77, *SE* = .29, *t* = -2.62, *p* = .009. Given that these results were not supported by the primary analyses, they should be interpreted with caution.

For proliferation in response to poly (I:C) stimulation, only the effect of childhood SES on the intercept (but not slope, *p* = .96) was significant, *β* = .22, *SE* = .10, *t* = 2.25, *p* = .03, with higher childhood SES predicting greater proliferation. Neither childhood unpredictability (*p*s > .51), nor adverse childhood experiences (*p*s > .68) predicted the intercept or slope. No interaction between sex and any childhood environmental variable reached significance (*p*s > .35).

**Phagocytosis**

As was found in the previous analyses, neither childhood unpredictability (*p* =.14) nor adverse childhood experiences (*p* = .19) significantly predicted phagocytosis. The interactions between these variables and sex, again, did not reach statistical significance (*p*s > .11). The interaction between childhood SES and sex remained significant, *β* = -.31, *SE* = .13, *t* = -2.39, *p* = .02, with higher childhood SES predicting greater phagocytosis in men, *β* = .29, *SE* = .13, *t* = 2.32 *p* = .02, but not women, *β* = -.17, *SE* = .14, *t* = -1.20, *p* = .23.

**Natural Killer Cell Cytotoxicity**

Results again revealed significant two-way interactions between childhood SES and sex predicting the intercept, *β* = -.56, *SE* = .28, *t* = -1.99, *p* = .047, linear slope, *β* = -1.34, *SE* = .27, *t* = -4.91, *p* < .001, and quadratic slope, *β* = 1.16, *SE* = .30, *t* = 3.88, *p* < .001. No other interactions were significant (*p*s > .10), nor were any main effects of childhood unpredictability (*p*s > .36) or adverse childhood experiences (*p*s > .25).

Unpacking the significant interaction revealed that, for men, higher childhood SES predicted greater NK cell cytotoxicity at the 12.5:1 E:T ratio, at a level of marginal significance (intercept), *β* = .20, *SE* = .11, *t* = 1.92, *p* = .055, and a greater increase in NK cell cytotoxicity as E:T ratios increased (linear slope), *β* = .40 *SE* = .10, *t* = 3.89, *p* < .001, with a diminishing effect at the highest E:T ratio (quadratic slope), *β* = -.36, *SE* = .12, *t* = -2.98, *p* = .003. In women, while childhood SES did not predict the intercept, *β* = -.12, *SE* = .12, *t* = -.99, *p* = .32, higher childhood SES predicted a reduced increase in NK cell cytotoxicity as E:T ratios increased, *β* = -.37, *SE* = .13, *t* = -2.96, *p* = .003, an effect which diminished at the highest E:T ratio, *β* = .32, *SE* = .13, *t* = 2.40, *p* = .02.

**PBMC Cytokine Release**

Results again revealed that there were no significant interactions between any childhood environmental measure and sex on either spontaneous cytokine release (*p*s > .22) or LPS-induced cytokine release (*p*s > .67). There were no main effects of any predictor on LPS-induced cytokine release (*p*s > .38). However, both childhood SES, *β* = -.33, *SE* = .09, *t* = -3.73, *p* < .001, and childhood unpredictability, *β* = -.29, *SE* = .10, *t* = -2.95, *p* = .003, emerged as significant predictors of spontaneous cytokine release. Specifically, higher childhood SES and higher unpredictability each independently predicted lower release of proinflammatory cytokines from PBMCs plated in media only. Adverse childhood experiences did not significantly predict spontaneous cytokine release (*p* = .71).

**Testosterone**

As in the primary analysis, there was a significant two-way interaction between childhood SES and sex on testosterone levels, *β* = .40, *SE* = .16, *t* = 2.50, *p* = .01. There were no significant main effects or interactions involved either childhood unpredictability (*p*s > .20) or adverse childhood experiences (*p*s > .53). Unpacking the interaction revealed that higher childhood SES, again, predicted lower levels of testosterone in men, *β* = -.25, *SE* = .07, *t* = -3.62, *p* < .001. In women, childhood SES did not significantly predict levels of testosterone, *β* = -.08, *SE* = .15, *t* = -.48, *p* = .63.

**Additional Discussion**

Interestingly, in addition to finding that childhood SES differentially impacted immune function within each sex, differences in immune function between the sexes also differed by level of SES and immune measure. For example, men with a higher childhood SES showed greater phagocytic ability and NK cell cytotoxicity than did women with a higher childhood SES, but there were no differences between the sexes at low childhood SES. Conversely, women generally tended to be higher than men in stimulated PBMC proliferation, regardless of childhood SES. Future research would benefit from delving more deeply into these differences to better elucidate the roles that sex and early life experiences each play in shaping the activities of the immune system.

One unanticipated result from the current research was that women’s NK cell cytotoxicity was negatively related to their early life SES. Though this pattern did not reach conventional levels of significance at most of the E:T ratios, that this relationship fell in the opposite direction of the relationship observed for men warrants comment. Although the reasons for this finding are unclear, there is some evidence that elevated NK cell activity may increase women’s risk of complications during pregnancy (6). Further, research finds that NK cell cytotoxicity varies across the ovulatory cycle, decreasing prior to ovulation (i.e., during the follicular phase of the cycle) compared to after ovulation has occurred (i.e., during the luteal phase; 7). Accordingly, it may be beneficial for women to downregulate NK cell cytotoxicity when conception becomes more likely. Given that all of the women in the current research were of reproductive age and participated during the follicular phase of their cycle, it is possible that women with a higher childhood SES are better able to temporarily suppress NK cell activity prior to ovulation than women with a lower childhood SES. Although speculative, such an explanation may lend insight into one potential facet of why women with a lower SES are at a greater risk for adverse pregnancy outcomes (compared to women with a higher SES; 8–9).

Another unexpected result was that lower childhood SES predicted greater spontaneous, but not LPS-stimulated, proinflammatory cytokine release by PBMCs for both men and women. The finding that early life stress increases the inflammatory response of PBMCs to mitogen stimulation is well-documented in the literature (e.g., 10–11). However, to our knowledge, this is the first research, to date, examining the effects of early life stress on the inflammatory activity of PBMCs a) while treating stimulated and unstimulated (i.e., spontaneous) cytokine release as separate measures and b) in the context of examining the impact of early life stress on a multiple additional measures of immune function. Regarding point *a*, although a number of studies have shown that measuring the unstimulated release of proinflammatory cytokines by PBMCs *in vitro* provides a useful index of basal inflammatory activity (e.g., 7–8), many studies that collect such data often do not analyze them. Instead, unstimulated cytokine release is only controlled for, or subtracted from stimulated cytokine release to compute a change score. Although each of these methods for analyzing *in vitro* cytokine release data may be entirely appropriate depending on one’s primary research question, the current findings suggest that how such data are to be analyzed should be carefully considered. Moreover, it is possible that significant relationships between early life stress (or any variable of interest) and the inflammatory activity of PBMCs in previous research may have been reduced or masked by failing to consider unstimulated and stimulated cytokine release as separate outcomes.

**Table S1.** Summary of Model Fit Indices

| **Model** | ***χ*^2^ (*df*)** | **CFI** | **RMSEA** | **SRMR** |
| --- | --- | --- | --- | --- |
| *Unconditional Models* |  |  |  |  |
| NK Cytotoxicity Model – Linear | 119.30 (6)*** | .79 | .36 | .15 |
| NK Cytotoxicity Model – Linear and Quadratic | 1.45 (2) | 1.00 | .00 | .01 |
| Proliferation Model – Media | 1.42 (1) | 1.00 | .06 | .01 |
| Proliferation Model – LPS | .13 (1) | 1.00 | .00 | .004 |
| Proliferation Model – PHA | .09 (1) | 1.00 | .00 | .007 |
| Proliferation Model – Poly (I:C) | 1.92 (1) | 1.00 | .08 | .02 |
| Cytokine Release Model | 6.88 (3) | .99 | .05 | .03 |
| *Final Models* |  |  |  |  |
| NK Cytotoxicity Model – Linear and Quadratic | 4.025 (5) | 1.00 | .00 | .01 |
| Proliferation Model – Media | 8.12 (4) | .98 | .08 | .03 |
| Proliferation Model – LPS | 7.09 (4) | .99 | .07 | .02 |
| Proliferation Model – PHA | 6.70 (4) | .98 | .07 | .04 |
| Proliferation Model – Poly (I:C) | 7.84 (4) | .98 | .08 | .04 |
| Cytokine Release Model | 7.47 (7) | .99 | .01 | .02 |

*Note*. Model fit for final models. CFI = comparative fit index; RMSEA = root mean square error of approximation; SRMR = standardized root mean square residual. **p* < .05.

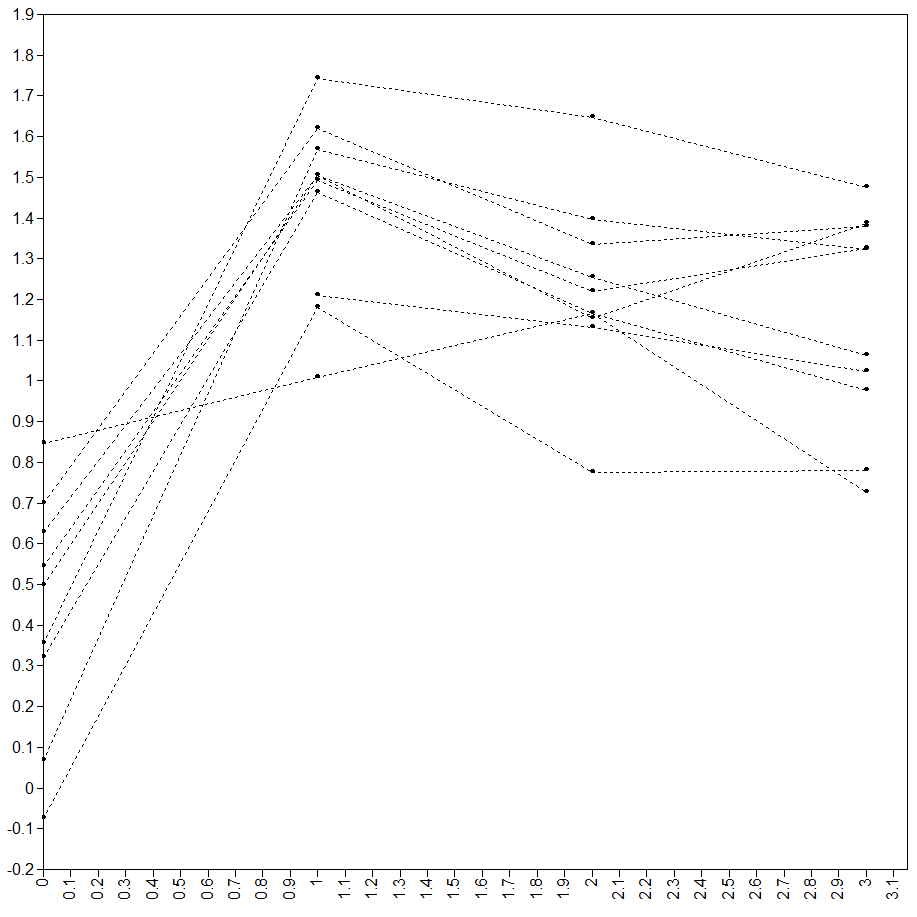

*Figure S1.* Interleukin-1 beta release from peripheral blood mononuclear cells over time in the absence of stimulation (i.e., spontaneous release) *in vitro*. Shown here are data from a random subset of 10 participants. Cytokine levels (pg/mL log-transformed) are shown on the Y axis, time is shown on the X axis.

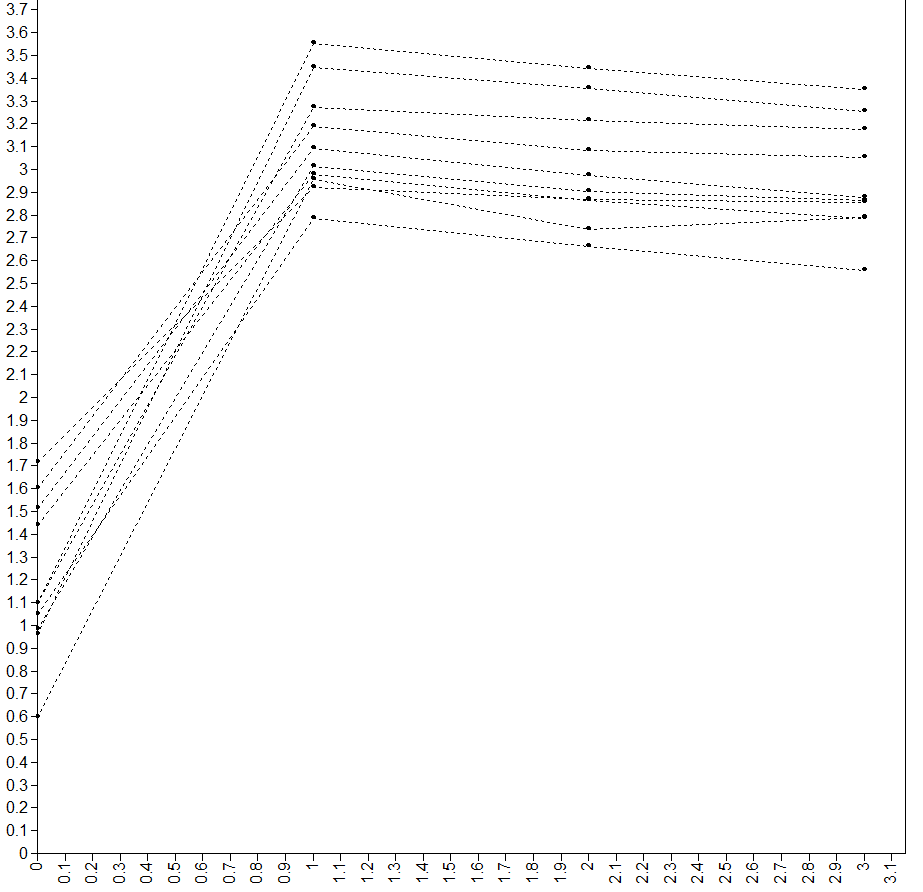

*Figure S2.* Interleukin-1 beta release from peripheral blood mononuclear cells after stimulation with lipopolysaccharide over time *in vitro*. Shown here are data from a random subset of 10 participants. Cytokine levels (pg/mL log-transformed) are shown on the Y axis, time is shown on the X axis.
